# Beyond the Label “Major Depressive Disorder” – Detailed Characterization of Study Population Matters for EEG-Biomarker Research

**DOI:** 10.1101/2025.03.17.25324119

**Authors:** Roman Mähler, Alexandra Reichenbach

## Abstract

Major Depressive Disorder (MDD) is a prevalent, multi-faceted psychiatric disorder influenced by a plethora of physiological and environmental factors. Neuroimaging biomarkers such as diagnosis support systems based on electroencephalography (EEG) recordings have the potential to substantially improve its diagnostic procedure. Research on these biomarkers, however, provides inconsistent findings regarding the robustness of specific markers. One potential source of these contradictions that is frequently neglected may arise from the variability in study populations. This study systematically reviews 66 original studies from the last five years that investigate resting-state EEG-biomarker for MDD detection or diagnosis. The study populations are compared regarding demographic factors, diagnostic procedures and medication, as well as neuropsychological characteristics. Furthermore, we investigate the impact these factors have on the biomarkers, if they were included in the analysis. Finally, we provide further insights into the impact of diagnostic choices and the heterogeneity of a study population based on exploratory analyses in two publicly available data sets. We find indeed a large variability in the study populations with respect to all factors included in the review. Furthermore, these factors are often neglected in analyses even though the studies that include them tend to find effects. In light of the variability in diagnostic procedures and heterogeneity in neuropsychological characteristics of the study populations, we advocate for more differentiated target variables in biomarker research then simply MDD and healthy control. Furthermore, the study populations need to be more extensively described and analyses need to include this information in order to provide comparable findings.

## 1 Introduction

Major depressive disorder (MDD) is a global health burden affecting all areas of life (WHO, 2017). Lack of interest and reduced drive over a longer period are its key characteristics. Beyond this, it is a rather heterogeneous disorder in terms of symptoms and disease progression (Bundesärztekammer, Kassenärztliche Bundesvereinigung, & Arbeitsgemeinschaft der Wissenschaftlichen Medizinischen Fachgesellschaften, 2022). MDD manifests in episodes with high recurrence (Marx et al., 2023) and can be accompanied by psychosis, anxiety, or cognitive dysfunction, among other symptom dimensions (Bundesärztekammer et al., 2022). All these different manifestations of the disorder are summarized under the diagnostic label *MDD*. Current clinical practice is to screen for MDD with neuropsychological questionnaires such as the PHQ-9 (Kroenke, Spitzer, & Williams, 2001; Marx et al., 2023) and diagnose it using semi-structured interviews based on DSM-5 or ICD-11 (Bundesärztekammer et al., 2022; Kołodziej, Magnuski, Ruban, & Brzezicka, 2021). However, those tools usually only reflect the patients’ current mood and diagnoses are influenced by the procedure and clinician’s experience, which makes them rather subjective (Cai et al., 2022). In order to support the diagnostic procedure with objective tools, imaging biomarkers based on recording abnormal brain structure or function associated with the disorder are investigated (Marx et al., 2023; Otte et al., 2016). Such biomarkers can also improve treatment decision and monitoring, and development and evaluation of therapies (Kupfer, Frank, & Phillips, 2012).

Electroencephalography (EEG) provides a non-invasive, easy to use, and low-cost tool to assess pathological alterations in brain physiology and is therefore an attractive choice for clinical application. However, contradictory findings regarding the usefulness of specific EEG biomarkers for MDD (Greco et al., 2021) and problems in reproducibility (Botvinik-Nezer & Wager, 2023; Van Dijk et al., 2022) demonstrate that there are gaps that need to be closed before those imaging biomarkers can be applied for diagnostic support. On the one hand, a plethora of signal processing and analysis choices impedes comparability and reproducibility across studies (Niso et al., 2022).

Recent reviews focus on the variety of biomarkers with potential diagnostic value that can be extracted from the EEG signal (Greco et al., 2021; Knociková & Petrásek, 2021) and the technical advancements in data processing and analysis algorithms (Dev et al., 2022; Yasin et al., 2021). On the other hand, there is the problem of small sample sizes that many researchers criticize in the light of the heterogeneity of MDD (Malgaroli, Calderon, & Bonanno, 2021). This problem is aggravated by the manifold of individual genetic, physiological, and environmental factors influencing MDD (Otte et al., 2016) and additionally for EEG biomarker research, the manifold of genetic (Bazanova & Vernon, 2014) and physiological (Brismar, 2007) factors influencing the EEG signal. These effects on the EEG signal might interact with the disorder, or be independent of it.

Demographic factors such as age and gender are known as mediating factors in MDD (Marx et al., 2023) as well as affecting the EEG signal (Polich, 1997; Polunina & Lefterova, 2012; Shearer, Cohn, Dustman, & LaMarche, 1984; Tröndle et al., 2023). Many more demographic, genetic, psychological, social, and behavioral factors influence MDD course (Marx et al., 2023) and treatment response (Kennis et al., 2020). Further factors known to affect the EEG signal include e.g. handedness (Papousek & Schulter, 1999) or current mental state such as fatigue or stress (Ismail & Karwowski, 2020; Tran, Craig, Craig, Chai, & Nguyen, 2020; Vanhollebeke et al., 2022). Central decisions in every clinical study are the diagnosis or operationalization of the target variable(s), and the definition of inclusion criteria for the clinical groups. This introduces another source of heterogeneity, since diagnosis of MDD is neither standardized nor objective. Even less defined is who qualifies as healthy control (HC) participant. Along this line, participants might have different expressions of symptoms, co-morbidities/other diseases, or take drugs of any kind. All these factors can influence both MDD course as well as EEG signals. However, the study populations on which research for EEG-biomarker for MDD has been conducted, has been neglected so far in the systematic search for the origins of heterogeneity in research findings.

This work aims to answer the question of comparability of study populations across current studies on EEG biomarker for MDD by providing a systematic overview about the variability in participants with regard to the aforementioned factors. Furthermore, we are interested whether and how information beyond the labels MDD and HC are included in the analysis and if it is, whether this can improve EEG biomarker research. Furthermore, in order to demonstrate the heterogeneity of study populations and the impact of diagnostic procedures, we complement the review with some exploratory analyses on publicly available data sets that are frequently used in MDD biomarker research.

## 2 Article Methods

### 2.1 Systematic review

The systematic review was conducted according to the PRISMA guidelines (Page et al., 2021). We focused on finding representative original studies on resting-state EEG biomarker research for unipolar depression, resp. MDD diagnosis or recognition rather than treatment in order to keep the use case concise. The search was conducted in the PubMed database on October 3^rd^ 2024 since we expect clinically relevant studies to be published in an outlet indexed in PubMed. The search string (‘major depressive disorder’ OR MDD) AND (biomarker OR diagnosis OR detection) AND (electroencephalography OR EEG) NOT (treatment OR TMS OR ‘transcranial magnetic stimulation’ OR sleep) was used with a filter on the last five years to include only recent research. Papers were excluded if they met at least one of the following exclusion criteria: (1) the aim of the paper was not diagnosis or detection of MDD; (2) MEG or other image modalities were used; (3) task-EEG, event-related potentials, or sleep EEG was recorded; (4) the purpose of the study was data augmentation; (5) the study contains only bipolar patients or a specific subgroup of MDD; (6) the study does not have a HC group; (7) the paper is a review; (8) the paper is not published in English or not accessible.

We extracted information about the three categories demographic data, diagnosis and medication, and neuropsychological tests to characterize the study populations as detailed as possible. Demographic data includes the size of the study population, age, gender, and ethnicity as smallest set of overlapping information across studies. Diagnosis includes the procedures to obtain the labels *MDD* and *HC* including exclusion criteria for participants. Regarding medication, we included all information on drugs available from the methods or results of the studies. Neuropsychological tests assess the severity of psychiatric diseases, symptom dimension, or cognitive function, usually with a (self-administered) questionnaire. They can be used for screening, complement diagnosis, or provide a more detailed description of a study population. For all three categories, we also extracted information about the consideration of these factors in the analysis and their impact. Since a substantial amount of studies is based on publicly available data sets, we separate the information of studies based on own data from studies based on public data.

### 2.2 Analysis of publicly available data sets

For further demonstration of the influence of the factors investigated in the review and the heterogeneity of a study population when considering further data about the participants, we conduct exploratory analyses on two publicly available data sets that are also used in some of the studies presented in this review (for details see chapter 3.2).

The CAV data set (Cavanagh, Bismark, Frank, & Allen, 2019) provides rich diagnostic information, including two different neuropsychological tests for the assessment of depression severity, which allows for the comparison of different diagnostic scenarios. The MODMA data set (Cai et al., 2022) provides six neuropsychological tests, which is well suited for an analysis of study population heterogeneity.

Group comparisons are conducted with analyses of variance (ANOVA), *t*-tests, or Fisher’s exact test dependent on the number of groups and nature of the data. Significance level is assumed with α<0.05, post-hoc tests are Bonferroni-corrected. Exploratory analyses for relationships between variables are investigated with Pearson’s correlation or general linear regression models. Exploratory patient stratification is conducted with *k*-means and hierarchical clustering with Euclidean distance based on min-max normalized data.

## 3 Results

Each result chapter from 3.3 onwards first describes the studies based on own data and subsequently the publicly available data sets, or the studies based on them, respectively. Moreover, first the key characteristics are described and summarized, and subsequently their consideration in the analysis of studies and their influence are reported.

### 3.1 Study selection

The initial database search yielded 193 papers. We excluded 91 papers during title screening and further 25 during abstract screening. From the remaining 77 papers, we excluded eleven more during full paper review. Reasons for exclusion were: Electrocardiography data was used instead of EEG (n=1), event-related potentials were analyzed and not resting-state EEG (n=3), no HC (n=2), not accessible (n=3), not in English (n=1). Additionally, one paper reported using the MODMA data set but since the data presented did not match this data at all, the study was excluded as well. This left 66 papers to include in the review, 34 of which collected own data, and 32 worked with publicly available data sets.

### 3.2 Publicly available data sets

The 32 studies working exclusively with public data utilized seven different data sets. The most frequently used data set was MODMA (Cai et al., 2022). Nine studies are based solely on this data (Deng, Fan, Lv, & Sun, 2022; W. Liu, Wang, Hamalainen, & Cong, 2022; B. Wang et al., 2023; W. Wu, Ma, Lian, Cai, & Zhao, 2022; B. Zhang et al., 2023; B. Zhang et al., 2021; J. Zhang, Xu, & Yin, 2023; Zhao, Gao, et al., 2022; Zhao, Pan, et al., 2022), five more studies additionally included other public data sets (Chu et al., 2024; Kabbara et al., 2022; Movahed, Jahromi, Shahyad, & Meftahi, 2022; X. Sun et al., 2024; Y. Wang, Zhao, et al., 2024), and another study replicated their findings based on own data with this data set (Soni, Seal, Yazidi, & Krejcar, 2022). MODMA is a multi-modal data set for MDD research containing three experiments: eyes-closed resting-state EEG data with three or 128 electrodes, for the latter one task data during EEG recording as well. The third experiment collected natural language but not EEG. Descriptions in the subsequent chapter are only for the 128-electrode resting-state data (n=24/29 MDD/HC), because this set was used most frequently in the studies of this review.

Nearly as frequently used is the data set collected from Mumtaz and colleagues (MUM) (Mumtaz, Xia, Mohd Yasin, Azhar Ali, & Malik, 2017). MUM was used in eleven studies alone (Ataei & Wang, 2022; Ellis, Sancho, Miller, & Calhoun, 2024; Ellis, Sattiraju, Miller, & Calhoun, 2023; M. Kang, Kwon, Park, Kang, & Lee, 2020; Khadidos, Alyoubi, Mahato, Khadidos, & Nandan Mohanty, 2023; Mahato & Paul, 2019; Movahed, Jahromi, Shahyad, & Meftahi, 2021; Saeedi, Saeedi, & Maghsoudi, 2020; Tang, Huang, Liu, & Yu, 2024; Zhou, Sun, Wang, & Jiang, 2024) and in three studies in combination with other data sets (L. Li, Wang, Li, & Zhao, 2024; Movahed et al., 2022; X. Sun et al., 2024). The public data set contains eyes-closed and eyes-open resting-state EEG data as well as task-EEG data with 19 electrodes from 34 MDD patients and 30 HC.

The data set provided by Cavanagh and colleagues (CAV) (Cavanagh et al., 2019) was used in four studies alone (Thoduparambil, Dominic, & Varghese, 2020; Trambaiolli & Biazoli, 2020; Yun & Jeong, 2021; Zandbagleh, Sanei, & Azami, 2024) and together with other data sets in five studies (Chu et al., 2024; Kabbara et al., 2022; L. Li et al., 2024; X. Sun et al., 2024; Y. Wang, Zhao, et al., 2024). The data set includes task as well as resting-state EEG data with 64 electrodes from 120 participants altogether.

The DRYAD data (Kołodziej et al., 2021) was used in one study in combination with other data sets (Chu et al., 2024) and consists of three small data sets: Nowowiejska (NOW; n=55), DiamSar (DIA; n=95), and Wronski (WRO; n=82). All three data sets contain 64-channel eyes-closed resting-state EEG data.

The data sets EMBARC (Webb et al., 2016), TDBRAIN (Van Dijk et al., 2022), and B-SNIP (Tamminga et al., 2017) were used by one study each (Ciarleglio, Petkova, & Harel, 2022; Gour et al., 2023; Lechner & Northoff, 2024). The EEG data of these three data sets are part of larger data collections. EMBARC was a multi-site drug study including resting-state EEG as well as fMRI data. TDBRAIN was collected over 20 years, containing EEG data of patients with different diagnoses, most frequent were MDD, attention deficit disorder, subjective memory complaints, and obsessive-compulsive disorder. B-SNIP primarily contains data from schizophrenia, schizoaffective disorder, or psychotic bipolar I disorder patients, and their direct relatives. The data set includes resting-state and task EEG, fMRI, and blood samples.

### 3.3 Study information and demographic data

More than half of the studies with own data collection were conducted in Asia, with more than half of the investigated subjects participating at an Asian location (Table 1). This information serves as proxy for ethnicity since most studies do not provide explicitly the ethnicity of their participants.

**Table 1.** Locations of data collection for studies with own data collection. ^a^1x Belgium; 1x Germany; 1x Netherlands; 1x Germany, Austria, & Luxembourg

| Location | Number of |  | References |
| --- | --- | --- | --- |
|  | *Studies*<br>*(n=34)* | *Participants*<br>*(n=31)* |  |
| China | 17 | 1692 | (Chen et al., 2024; Hong et al., 2021; Y. Huang et al., 2023; X. Kang et al., 2024; Lei, Chen, Li, Zhou, & Cui, 2023; Y. Li et al., 2022; H. Lin et al., 2022; Z. Lin et al., 2020; S. Liu et al., 2022; W. Liu et al., 2020; Shao et al., 2021; S. Sun et al., 2023; Teng et al., 2022; Y. Wang, Peng, et al., 2024; Z. Wu et al., 2022; Xie et al., 2024; Xue et al., 2024) |
| South Korea | 3 | 272 | (Choi et al., 2021; Jang, Lee, Lee, Huh, & Chae, 2020; Shim, Hwang, & Lee, 2023) |
| Taiwan | 3 | 717 | (Duncan et al., 2020; Huang, Fan, & Lin, 2023; C. T. Wu et al., 2021) |
| Russia | 2 | 143 | (Mitiureva et al., 2024; Proshina, Martynova, Portnova, Khayrullina, & Sysoeva, 2024) |
| Europe<sup>a</sup> | 4 | 613 | (Benschop et al., 2022; Périard et al., 2024; Sarisik et al., 2024; Sverdlov et al., 2021) |
| USA | 3 | 544 | (Lord & Allen, 2023; Murphy et al., 2020; Umemoto et al., 2021) |
| Australia | 1 | 100 | (Sharpley, Bitsika, Shadli, Jesulola, & Agnew, 2023) |
| unknown | 1 | 33 | (Soni et al., 2022) |

Since two data sets were used twice each and one study did not provide the number of participant, the number of participants is based on 31 studies. The publicly available data sets were collected in China (MODMA), Malaysia (MUM), Poland (NOW, DIA, WRO), Netherlands (TDBRAIN), and USA (CAV, EMBARC, B-SNIP).

Half of the studies base their results on 80 or less participants total (**Fehler! Verweisquelle konnte nicht gefunden werden.**A) with most of the studies having about an even split between the clinical groups (Fig. 1B, top box). Studies tend to include more female than male participants in both clinical populations (Fig. 1B, bottom boxes) with two studies investigating female participants only (Shim et al., 2023; Umemoto et al., 2021).

**Fig. 1.**
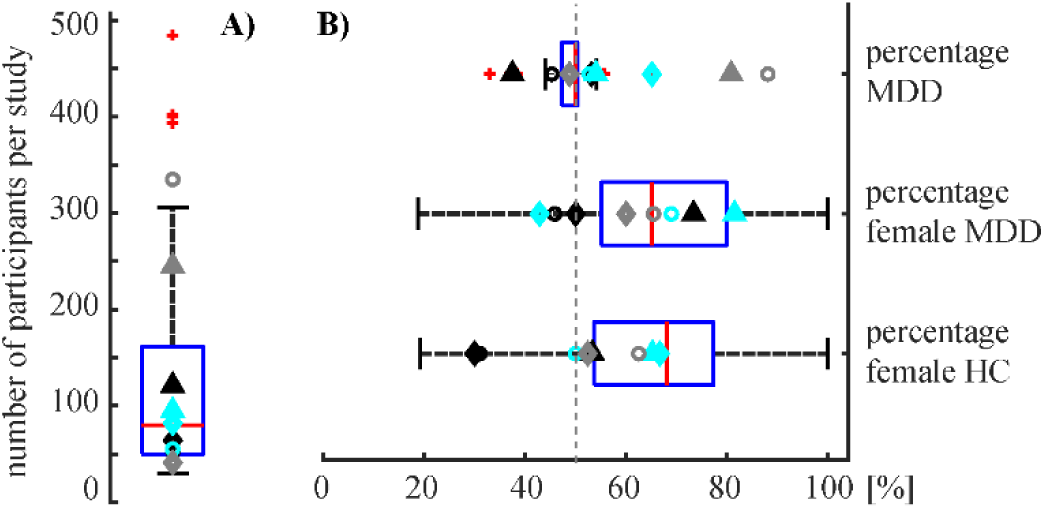
Basic characteristics of the studies. A) Overall number of participants for studies with own data (boxplot: n=31) and overlaid the population sizes of the public data sets. B) Percentages of MDD patients relative to the sum of MDD patients and HC (boxplot: n=31) and the percentages of female participants in the two clinical sub-groups (boxplots: n=27) for the studies with own data. Overlaid are the respective information for the public data sets. Black symbols: MODMA (circle), CAV (diagnosis based on BDI; triangle), MUM (diamond); cyan symbols: NOW (diagnosis based on diagnosis; circle), DIA (A: all; B: diagnosis based on diagnosis and BDI score (n=50); triangle), WRO (A: all; B: diagnosis based on BDI score (n=86); diamond); grey symbols: EMBARC (circle), TDBRAIN (triangle), B-SNIP (diamond). The first six data points are based on the public data itself, the latter three are based on the studies included in the review.

Age distributions of studies vary widely (Fig. 2). The older the study population average the higher the spread in the population, except for one study with a rather old but confined age distribution (Z. Wu et al., 2022). One study specifically recruited adolescents up to 18 years (Umemoto et al., 2021), all other studies targeted adults.

**Fig. 2.**
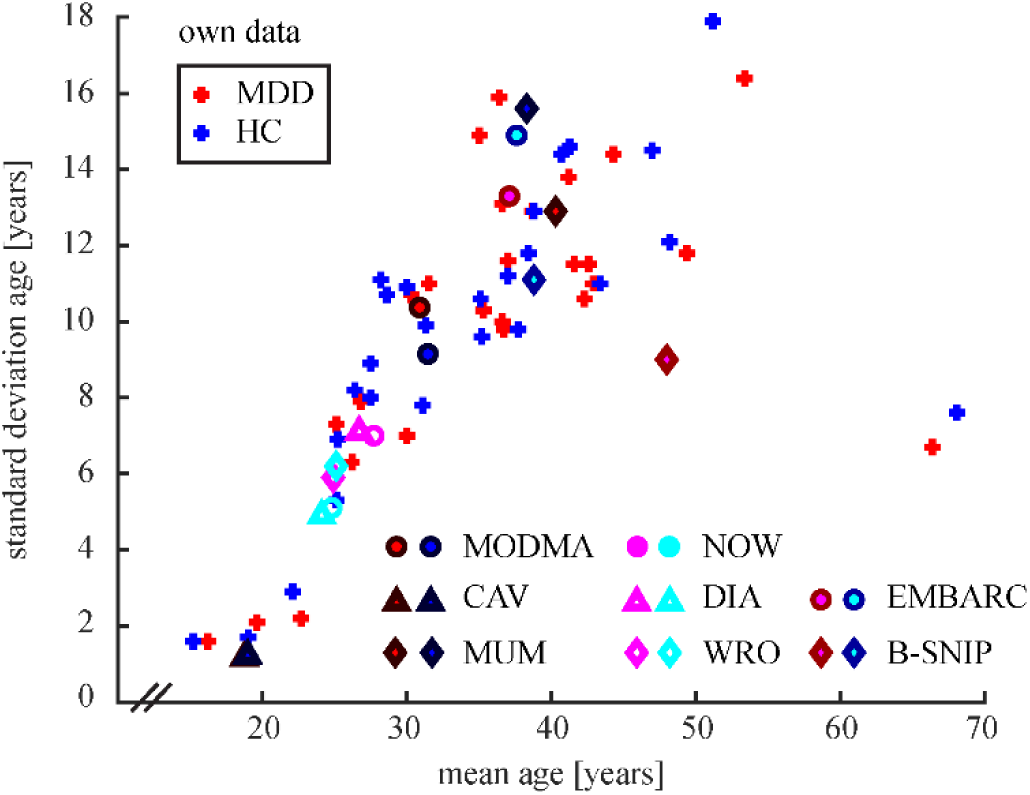
Age distributions of participants in studies with own data collection (n=26) and in public data sets. Color coding for the publicly available data sets: reddish colors mark MDD patients, blueish colors mark HC. Diagnosis is defined analogue to Fig. 1B. For study abbreviations see chapter 3.3.

Seven studies based on own data considered gender or age in their analysis. Two studies included gender: Lord and Allen (2023) conducted additional analyses for gender separately and found differences in EEG complexity metrics between those groups. Three studies (Benschop et al., 2022; Jang et al., 2020; Mitiureva et al., 2024) used age and gender as covariates in statistical comparison of clinical groups or regressed them out in correlation analyses. Two more studies treat only age as covariate (Périard et al., 2024; Umemoto et al., 2021); note that the latter one included only female participants. One study specifically investigated pathological aging processes and found that aging affects the diagnostic capability of EEG biomarkers (Sarisik et al., 2024).

The studies using only MODMA, CAV, or MUM data did not consider gender, age, or any other demographic information in their analysis. One study using both MOD and CAV did not find an age but a gender influence in their regression models (Kabbara et al., 2022). The two studies using EMBARC (Ciarleglio et al., 2022) and B-SNIP (Lechner & Northoff, 2024) used age and gender as covariates in statistical comparison of clinical groups.

To summarize, the majority of studies is based on small sample sizes (n<100 per diagnostic group) with a disproportional high ratio of Asian participants. Studies are rather variable in their gender ratio and age distributions, both for studies based on own data as well as public data sets. Only few studies consider gender or age in the analysis. Those explicitly investigating the influence of these factors, however, tended to find one.

### 3.4 Diagnostic and medication information

All studies with own data separated their participants into the groups *MDD* and *HC*. This diagnostic label was used for classification in twelve studies, for group comparisons in 17 studies, and for both analysis approaches in five studies. Twenty-four of the studies based on public data used the diagnostic groups *MDD* and *HC* for classification, four for group comparisons, and two for both. One study based on the CAV data set (Trambaiolli & Biazoli, 2020) did not use that dichotomous diagnostic label at all but utilized neuropsychological scores as only target variable instead.

#### 3.4.1 Studies based on own data

For inclusion in the MDD group, 21 studies collecting their own data report the involvement of a psychiatrist or similarly trained clinician, five studies even confirmed the diagnosis with a second specialist. For inclusion in the HC group, only seven studies involved such a specialist. The use of a structured interview (SCID (First, 1997) or MINI (Sheehan et al., 1997)) is mentioned for the diagnosis of MDD in ten studies but only in five studies for the confirmation of inclusion in the HC group. The MDD diagnoses were based either on the Diagnostic and Statistical Manual of Mental Disorders (DSM) version 4 (Bell, 1994) or 5 (American Psychiatric Association & American Psychiatric Association, 2013) (n=8/11) or on the International Statistical Classification of Diseases and Related Health Problems (ICD) version 10 (Organization, 1992) or 11 (Reed et al., 2019) (n=4/1). Status of HC was confirmed by the DSM-4 or 5 criteria in 3/1 studies and by the ICD-10 or 11 criteria in one study each.

Five and one studies additionally used the Hamilton Depression Scale with 17 items (HAMD-17) (Hamilton, 1960) or Beck Depression Inventory Version 2 (BDI-II) scores (Beck, Steer, & Brown, 1996) (see chapter 3.5), respectively, as inclusion criterion in the MDD group. Three of those studies additionally used the Young Mania Rating Scale (YMRS) (Young, Biggs, Ziegler, & Meyer, 2000) for exclusion of bipolar patients. One study each confirmed HC status based on HAMD-17 or BDI-II. Two studies did not use their diagnoses but relied on BDI-II scores for grouping of participants into MDD and HC instead. Two other studies used the HAMD-17 or Self-Rating Depression Scale (SDS) scores instead of a diagnostic procedure for the MDD group, and eight for the HC group with the use of BDI-II (n=4), HAMD-17 (n=3), or SDS (n=1). Noteworthy, cutoff criteria on the scores for grouping differed across studies, even for the same test.

Two studies relied on self-report for inclusion in the MDD group, five for inclusion in the HC group. One study did not report any assessment criteria for inclusion in the MDD group, eight studies neglected this for the HC group.

General exclusion criteria for both groups were in seven studies intelligence or education below average, or learning disorder, pregnancy in four studies, and non-right-handedness in two studies.

Thirteen studies excluded MDD patients with any axis-I disorder co-morbidity except for anxiety disorder in two studies. HC were excluded if they had any axis-I disorder in eleven studies, or if they had a family history of psychiatric disorders in four studies. Nineteen or 18 studies excluded MDD patients or HC, respectively, if they had any neurological disorder, or some specific neurological disorders. Any other disease was an exclusion criterion for MDD or HC in four or five studies, respectively. Six or one study excluded MDD patients or HC if they had recently electroconvulsive or TMS therapy. One study excluded patients that were diagnosed with MDD as a secondary disorder next to, e.g. Parkinson’s disease. Finally, four studies included only first-episode MDD patients.

In one study, all MDD patients were medicated, in eleven studies some of the patients were medicated. Details about the medication varied from frequencies of specific drugs taken, descriptive group statistics about the active substances to simply the mention of patients taking anti-depressants. One study each excluded patients on lithium or tricyclic anti-depressants even if they allowed medication of patients otherwise. In nine studies, the MDD patients were drug-naïve or drug-free, in the latter case three studies mentioned since when they were at least drug-free. For the HC group, twelve studies report drug-free participants with one study confirming this with a drug test.

Furthermore, twelve or 14 studies excluded MDD patients or HC, respectively, when they had a history of alcohol or drug abuse.

#### 3.4.2 Publicly available data sets

For the MODMA data set, MDD patients were diagnosed with the MINI based on DSM-4 criteria and had to have a PHQ-9 (Patient Health Questionaire, chapter 3.5) score above five. Any participant without higher education or pregnant was excluded. MDD patients were excluded when they had any other axis-I disorder, were suicidal, or had brain damage. HC were excluded when they had a family history of psychiatric disorders. MDD patients were medication free for at least two weeks and an exclusion criterion for all participants was any other drug or psychotropic substance use.

For the MUM data set, MDD patients were diagnosed based on DSM-4 criteria, and had to have no other psychiatric or cognitive disorders, no epilepsy, and were not pregnant. HC were excluded when they had any axis-I disorder. MDD patients had to be medication free for at least two weeks and not abuse any drugs.

For the CAV data set, participants were first administered the BDI test and only participants with a score ≥13 were subsequently diagnosed with the SCID based on DSM-4 criteria. Based on the diagnosis, four groups were identified: current MDD, past MDD, no MDD, and not interviewed. HC had to have a BDI score below seven and no self-reported history of any axis-I disorder. Exclusion criteria for all participants was a history of trauma or seizure, or use of any psychoactive medication.

For the three DRYAD data sets, common diagnostic classifications and exclusion criteria were used. The MDD groups were diagnosed with the MINI based on ICD-10 criteria (NOW, DIA), all other groups were solely based on the BDI. HC needed to have a BDI-score ≤5 (NOW, DIA, WRO), subclinical participants were not formally diagnosed but had a score ≥10 (DIA, WRO), and the unclassified group had a BDI-score between five and ten. All participants were without neurological disorders or head injuries, and medication free.

#### 3.4.3 Consideration in analysis

Apart from the diagnostic labels *MDD* and *HC*, five studies with own data used additional diagnostic information to stratify the patient group, or characterize it further. Inclusion of the differentiation between current and remitted MDD showed that microstates between the two MDD groups are more similar to each other than to the HC group, but that small differences are also identifiable between the two MDD groups (Murphy et al., 2020). However, Lord and Allen (2023) did not find the expected differences between those two MDD groups in complexity metrics. In contrast, psychotic and non-psychotic MDD patients exhibit differential functional connectivity (Chen et al., 2024; Lei et al., 2023). Finally, Benschop et al. (2022) correlated age of onset, duration of the current episode, and number of total episodes with functional connectivity markers and found a substantial influence of the latter on some of their markers. Two studies investigated whether medicated and un-medicated patients differ in their EEG-markers but did not find differences (Sarisik et al., 2024; Umemoto et al., 2021). In contrast, the study based on the B-SNIP data set found a sign. correlation between medication load and their phase dynamic marker, and therefore used medication as covariate in their analysis (Lechner & Northoff, 2024).

To summarize, most studies base the inclusion in the MDD group on clinical standard diagnoses. However, procedures vary. Inclusion in the HC group was much less controlled. Neuropsychological tests sometimes supplemented the diagnosis or infrequently determined the diagnostic label. Most common exclusion criterion was the presence of a neurological disorder for both groups. Other exclusion criteria varied widely from none to a long list. The use of medication was also handled very differently. In the case of MDD, patients were most frequently either medication-free or the medication was reported. For HC, this information was rarely reported. Very few studies used additional diagnostic information in their analysis but those who did, found effects. The three studies that have examined medication report contradictory results.

### 3.5 Neuropsychological tests

Neuropsychological tests assess the severity of psychiatric diseases, symptom dimension, or cognitive function, usually with a (self-administered) questionnaire. They can be used for screening, complement diagnosis, and provide a more detailed description of a study population.

Five neuropsychological tests that operationalize depression severity are administered most frequently (Table 2): Hamilton Depression Scale with 17 items (HAMD-17) (Hamilton, 1960), Beck Depression Inventory (BDI) (Beck, Ward, Mendelson, Mock, & Erbaugh, 1961), currently most frequently used in version 2 (BDI-II) (Beck et al., 1996), 9-item subscale for depression from the Patient Health Questionnaire (PHQ-9) (Kroenke et al., 2001), Self-Rating Depression Scale (SDS) (Zung, 1965), and Montgomery-Åsberg Depression Rating Scale (MADRS) (Montgomery & Åsberg, 1979). Note that a clinician administers the HAMD-17 and MADRS while the other three tests are usually self-administered. Tests that operationalize anxiety sometimes complement the depression score, most frequently the Hamilton Anxiety Rating Scale (HAM-A) (Hamilton, 1969), and the State-Trait Anxiety Inventory (STAI) (Spielberger, Gonzalez-Reigosa, Martinez-Urrutia, Natalicio, & Natalicio, 1971) used in four and one study, respectively, the latter also in the CAV data set. Further neuropsychological tests that occurred more than once were the Young Mania Rating Scale (YMRS) (Young et al., 2000) and the Mini Mental State Examination (MMSE) (Folstein, Folstein, & McHugh, 1975) with four and two studies administering them, respectively.

**Table 2.** Most frequently administered neuropsychological tests for depression severity rating with their frequency in included studies and their severity rating ranges. Public data sets are only counted when they provide these scores with their data sets. For abbreviations see text. ^a^first row based on (Bundesärztekammer et al., 2022), second on values commonly used in research studies; ^b^for one study each, the version of the BDI is unknown.

| Test | Number (public) | None / Minimal | Mild | Moderate | Severe |
| --- | --- | --- | --- | --- | --- |
| HAMD-17 | 15 (1) | 0-8a<br>0-9 | 9-16<br>10-20 | 17-24<br>21-30 | 25-51<br>30-51 |
| BDI | 1<sup>b</sup> (3<sup>b</sup>) | 0-9 | 10-19 | 20-30 | 30-63 |
| BDI-II | 9 (2) | 0-12a<br>0-8 / 9-13 | 13-19<br>14-19 | 20-28<br>20-28 | 29-63<br>29-63 |
| PHQ-9 | 4 (1) | 0-4 / 5-9 | 10-14 | 15-19 | 20-27 |
| SDS | 4 | 25-49 | 50-59 | 60-69 | 70-100 |
| MADRS | 2 | 0-12 | 13-21 | 22-28 | 29-60 |

Distributions of the two most frequently used tests demonstrate a high variability across study populations (Fig. 3) with distinctive differences between diagnostic groups. Noteworthy, sometimes the tests are only administered to the MDD but not the HC group, as apparent for the HAMD-17 (Fig. 3A).

**Fig. 3.**
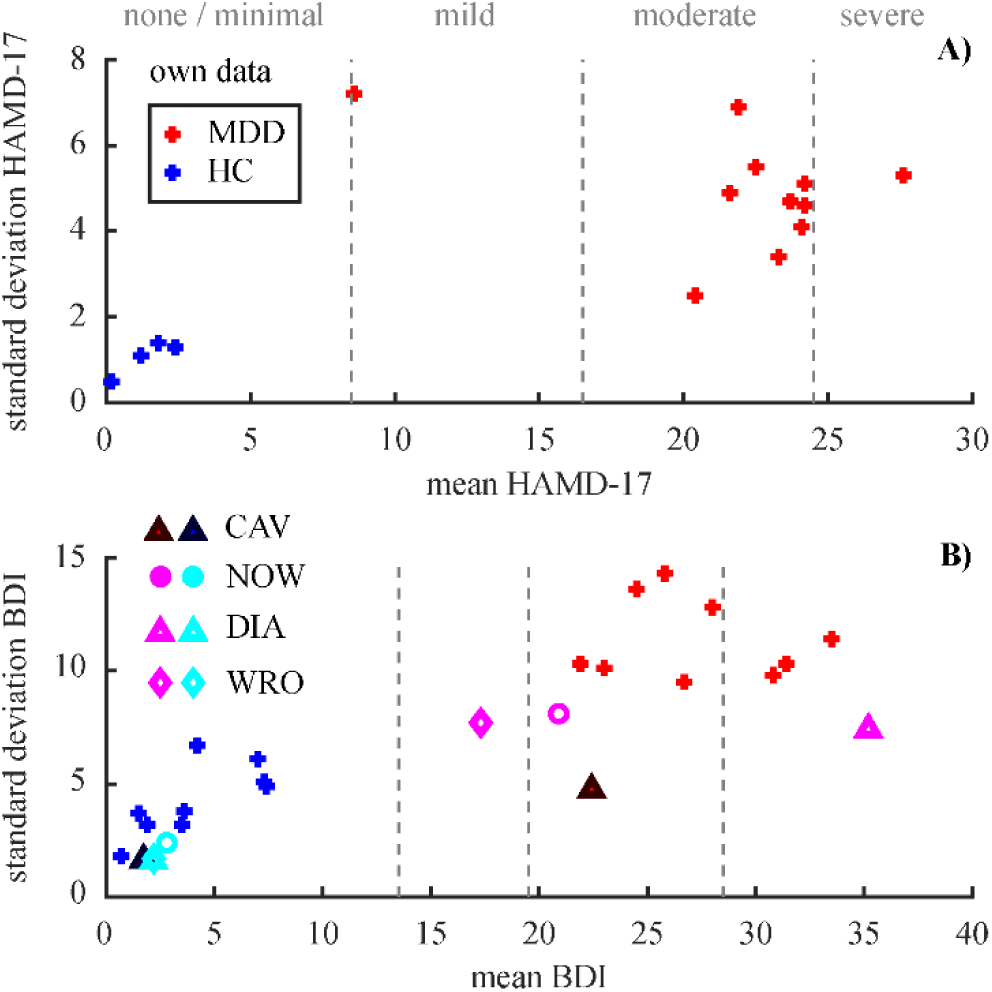
Distributions of the two most frequently used neuropsychological tests. A) HAMD-17 (n=11/4 for MDD/HC) B) BDI (n=9/9 for own data), both versions of the test. The dashed lines demarcate the cut-offs between severity categories for each of the tests. Color-coding and definition of diagnosis groups analogue to Fig. 2.

Half of the studies using own data included the test scores from at least one neuropsychological test in their analysis (Table 3). From the studies pooling data across both groups, five used the BDI-II score to operationalize the severity of depression, the PHQ-9, SRS, and MADRS were used once each. From the studies conducting the analysis on the MDD group only, six used the HAMD-17 and two the BDI-II. Six studies that use public data included the test score of at least one neuropsychological test (Table 3, marked with *). One of these studies, however, shows PHQ-9 values in their graphs that are actually not contained in the public data set (B. Zhang et al., 2021). The spectral markers seem to be rather contradictory in correlation with depression severity or other neuropsychological test scores. Connectivity markers, however, seem to be better correlated.

**Table 3.**
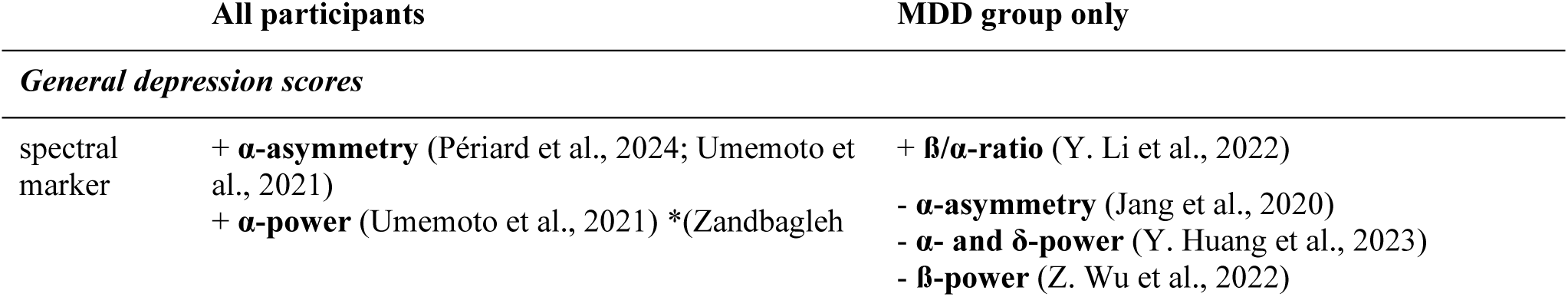
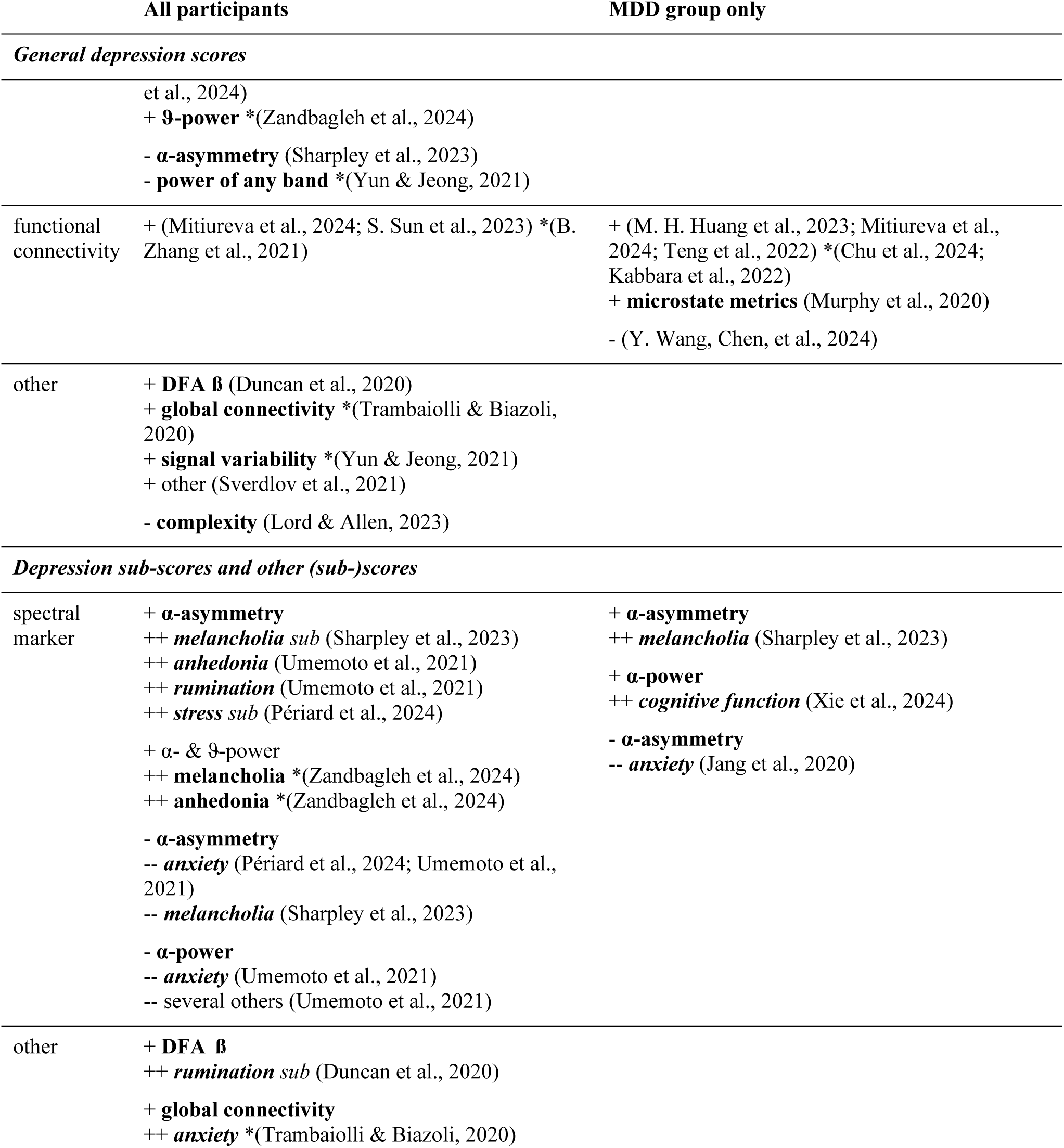
Summary of correlations with or regressions on neuropsychological test scores. The studies are pooled across individual tests. + denotes sign. correlation; – denotes reported n.s. correlation (note that not all n.s. correlations are listed due to publication bias). * denotes studies based on public data. Abbreviation: DFA: detrended fluctuation analysis.

In summary, the studies employed a variety of neuropsychological questionnaires, the most commonly used being the BDI and HAMD-17. These questionnaires categorize MDD into different levels of severity. However, the distribution of these scores varies greatly across studies. About half of the studies with own data also use the scores of neuropsychological questionnaires in their analysis, studies based on public data employ this practice less frequently. The results of the studies are partially inconsistent, especially regarding spectral markers.

### 3.6 Additional analyses on public data sets

The participants of the CAV data set can be divided into five groups based on their diagnostic procedure (Fig. 4A): 1) MDD_current_: diagnosis & BDI ≥13 (n=11; 54.5% female); 2) MDD_remitted_: diagnosis & BDI ≥13 (n=12; 75.0% female); 3) highBDI_noMDD_: diagnosis & BDI ≥13 (n=9; 66.7% female); 4) highBDI_undiag_: no diagnosis & BDI ≥13 (n=14; 92.9% female); 5) lowBDI: BDI ≤7 (n=75; 53.3% female).

**Fig. 4.**
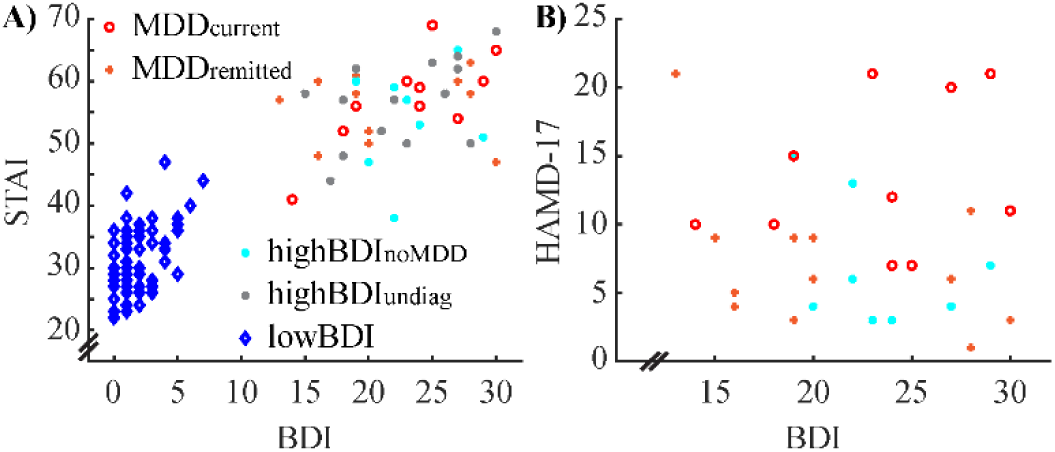
Relationship between the BDI and the STAI (A) and the BDI and the HAMD-17 scores (B) for the CAV data set. Grouping of participants is based on the diagnostic procedure (see text).

The first four groups have on average a moderate BDI-score (Fig. 4A; 22.2±4.9) without sign. differences between groups (*F*_3,42_=0.398; *p*=0.775). The HAMD-17 score, however, differs sign. between the first three groups (Fig. 4B; *F*_2,29_=5.169; *p*=0.012), with the MDD_current_ group (13.1±5.3) exceeding sign. both the MDD_remitted_ (7.3±5.3; *t*_21_=2.637; *p*=0.015; corr. *p*=0.045) and the highBDI_noMDD_ group (6.8±4.4; *t*_18_=2.852; *p*=0.011; corr. *p*=0.033). Note that only diagnosed participants were assessed with the HAMD-17 test.

Several dichotomous diagnosis groups (MDD/HC) can be formed from these five groups, depending on the research question. This practice been used in the studies based on this data set. 1) The strictest separation includes only MDD_current_ for the MDD group (n=11) and lowBDI for the HC group (n=75). 2) The second approach expands the MDD group by the MDD_remitted_ group (n=23) and keeps the HC group the same. 3) The diagnosis-agnostic approach groups participants solely based on BDI, e.g. BDI ≥13 for the MDD group (n=46) and BDI ≤7 for the HC group (n=75). The cutoff value, however, is of minor importance in this data set since there is a gap in the scores between eight and twelve. For a data set with continuous BDI scores, however, different cutoff values might be used and are used, e.g. group splits at BDI scores of nine or 13. 4) Only diagnosed MDD (current & remitted; n=23) are included in the MDD group while the HC group comprises all other participants (n=98). The last approach seems counterintuitive for the given data set. However, since a substantial amount of studies comprises ill-defined HC groups, this approach mirrors this practice.

Obviously, the distribution of BDI scores differs slightly between groups of these different “diagnosis” approaches. Furthermore, we find a sign. difference for gender between diagnostic groups in approach three only (odds ratio=2.479; *p*=0.034). This means that here either gender needs to be considered as a potential confound in the analysis, or e.g. the HC group is subsampled to match the MDD group – an approach that is possible here since the HC group is substantial larger than the MDD group. For none of the approaches we find a sign. age difference between groups (all *p*≥0.3).

The participants of the MODMA data set are usually grouped based on the diagnostic label given, which leads to a MDD group with 24 participants (45.8% female) and a HC group with 29 participants (31.0% female). Neither age nor gender differs sign. between these groups (both *p*>0.3). The education, however, is a potential confound with a sign. difference between groups (*t*_51_=-3.209; *p*=0.002), confirmed by a moderate but highly sign. correlation between education and PHQ-9 score (*r*=-0.45; *p*<0.001).

The exploratory stratification analysis on the MODMA data set is restricted to three scores, since more than three dimensions are not as intuitively to visualize. We chose the depression severity (PHQ-9) along with the anxiety (7-item Generalized Anxiety Disorder scale: GAD-7) (Spitzer, Kroenke, Williams, & Lowe, 2006) and sleep score (Pittsburgh Sleep Quality Index: PSQI) (Buysse, Reynolds, Monk, Berman, & Kupfer, 1989) because these two correspond to possible symptoms of depression (American Psychiatric Association & American Psychiatric Association, 2013; Malgaroli et al., 2021) as well as to disorders on their own right that can be co-morbid with MDD (X. Liu et al., 2007; Meng & Wang, 2023; Sevillano-Garcia, Manso-Calderon, & Cacabelos-Perez, 2007; Staner, 2010; Thaipisuttikul, Ittasakul, Waleeprakhon, Wisajun, & Jullagate, 2014; Zimmerman, Chelminski, & McDermut, 2002).

Visual exploration of the clustering hierarchy revealed four clusters that adequately separate the participants in one HC group and three MDD groups (Fig. 5A & B). For the latter, a small group of patients with low anxiety but sleeps problems (violet markers) stands out. The separation of the remaining MDD patients seems to be based on all three scores in a similar fashion. Based on the four clusters found in the hierarchical approach, the *k*-means algorithm was parametrized with four clusters as well. This algorithm separates the HC group as well and seems to be mainly driven by the PSQI/GAD-7 combination rather than by the PHQ-9 score (Fig. 5C & D).

**Fig. 5.**
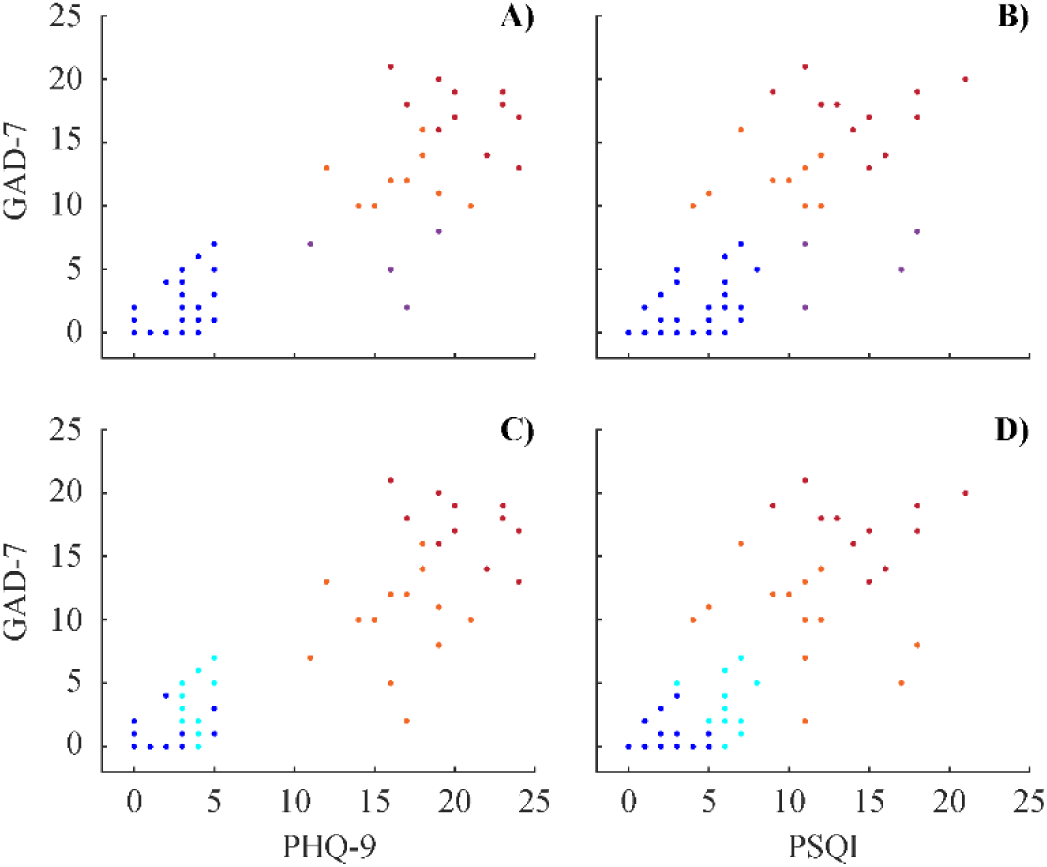
Clustering the MODMA data set based on depression (PHQ-9), anxiety (GAD-7), and sleep (PSQI) scores with hierarchical clustering (upper row) and k-means clustering (lower row). Colors in each row correspond to the same groups but colors across rows cannot be interpreted.

To summarize, grouping MDD and HC according to different criteria demonstrates that apart from a change in the distribution of MDD severity rating in the diagnostic groups, also the distribution of other variables like demographic information may shift, which might lead to potential confounders dependent on grouping. We also demonstrate that different neuropsychological tests like HAMD-17 and BDI can have different capabilities of separating between diagnostic subgroups. Moreover, adding further neuropsychological tests related to symptoms or co-morbidities such as sleep problems or anxiety, we show that already rather small and confined samples might contain clinically relevant subgroups.

## 4 Discussion

In a set of 66 original papers on EEG-biomarker research for MDD diagnosis from the last five years, we find a rather large diversity in population characteristics and differences in the definition of diagnostic groups across studies. This variety is a factor that contributes to the conflicting findings across studies are not surprising and characteristics of the study population needs to be taken into account when comparing study results or compiling meta-analyses, which is often overlooked. Our complementary exploratory analyses further demonstrate exemplary the influence of diagnostic procedure and choice of testing, as well as the heterogeneity in study samples.

A re-occurring criticism in clinical neuroimaging research is that findings are often based on rather small sample sizes (Botvinik-Nezer & Wager, 2023; Dev et al., 2022; Rakic, Cabezas, Kushibar, Oliver, & Llado, 2020). Our results confirm this shortcoming in recent research. We find only five studies collecting own data that have more than 300 participants, plus the EMBARC data as public data set. In contrast, 75% of the studies with own data included less than 160 participants. The two most frequently used public data sets that are used solo by 20 studies are rather small with 53 and 64 participants. While six studies replicate their findings across several public data sets, none of them pools the data together, and from the studies collecting own data, only one replicates their findings with public data. The latter one is a practice that can at least partially overcome the problem of developing a diagnostic procedure tailored only to a small, specific data set (Botvinik-Nezer & Wager, 2023).

The studies include on average a higher proportion of female participants, which mirrors the higher prevalence of MDD in women than men (Seedat et al., 2009). Even though female gender is considered a risk factor for MDD (Marx et al., 2023), and gender influences EEG signals (Polunina & Lefterova, 2012; Shearer et al., 1984), it was rarely included in the analyses. The two studies explicitly investigating a gender effect found one. Except for one study targeting adolescents, all studies collected data from adults, with a rather high variability in mean age and variance. Age is known to influence the EEG signal (Polich, 1997; Tröndle et al., 2023) and one study that specifically investigated the effect of age on EEG-biomarker for MDD found that MDD is diagnosed better in younger participants. Most patients experience their first episode in early adulthood (Kessler & Bromet, 2013), therefore this age group should be the main target for a diagnostic use case. However, due to the still prevalent social stigma the disorder is afflicted with (Stuart, 2016), there is likely a substantial number of older people still undiagnosed, who should also not be neglected in a diagnostic scenario.

Gender and age are only the most commonly reported variables describing the participants. Many more are known to influence MDD (see e.g. Marx et al. (2023)). Some of the studies assessed e.g. ethnicity, intelligence, or education. We found that education is a possible confound in the MODMA data set. None of the studies using this data, however, did mention the consideration of this variable in any analysis. Other studies used factors such as (low) intelligence and education, pregnancy, or (left-)handedness as exclusion criteria. Our results demonstrate that collecting additional information about the research participants and including them into analyses can improve the preciseness of the results and contribute towards resolving conflicting findings. This practice should therefore be increasingly used in biomarker studies.

All studies but one rely on the diagnostic labels MDD and HC for their main analysis. However, the definition of neither group is consistent across studies. A number of rather obvious reasons contribute to the element that the labels cannot be correct or clean. Any diagnostic procedure is subjective and diagnostic criteria changed over the years (American Psychiatric Association & American Psychiatric Association, 2013; Bell, 1994; Organization, 1992; Reed et al., 2019), MDD is a multifaceted disorder with several dimensions that can vary inter-individually in severity and current status (Malgaroli et al., 2021), and HC is per se an ill-defined group. There are also a number of less obvious reasons, such as the variability in operationalization of MDD severity by using different neuropsychological tests, or even when the same test is applied, different cut-off criteria are utilized. Our analysis on the CAV data demonstrates some possible definitions of diagnostic groups and the impact the groupings have. This suggests that instead of artificially creating two groups, the target variable for analysis should rather be continuous. However, we also show that dependent on whether the BDI or HAMD-17 score is considered, participants fall into different severity classes. Taken together with the exploratory analysis on the MODMA data where subgroups emerge when additional test scores are added, we rather recommend a multi-dimensional target variable rather than a one-dimensional severity scale. Scores of neuropsychological tests have indeed been used in additional analyses more than any other information, either depression severity rating, sub-scores of those scales, anxiety, or cognitive scores. The findings were mixed. However, given the design of the data collection in most studies, an artificial gap in MDD severity is introduced with rather high scores for the MDD group and rather low scores for the HC group. Data collection for continuous target variables should take care to obtain a more evenly distribution of this variable. In line with the recommendation for collecting and using more demographic information, the same applies to information about disease details and more meticulous description via neuropsychological tests.

Most variability across studies is introduced with the exclusion criteria and the drug status of participants. Some studies took great care to isolate the MDD diagnosis as tightly as possible in patients and exclude the possibility of any (psychiatric) disorders in the HC participants including genetic disposition. Other studies were completely oblivious about constraints for their participants, and anything in between. A similar picture is apparent regarding medication and drug use. Some studies take great care to establish that their participants, including the HC, are drug-free, or they report detailed medication-use. Other studies do not report anything about drug status. Very few studies consider details like (recent) nicotine or coffee consumption, even though these substances also affect the EEG signal (Conrin, 1980; Edwards & Warburton, 1982; Hammond, 2003; Norton, Brown, & Howard, 1992). In a diagnostic scenario, a person would possibly also have an additional disease unrelated to MDD or be on regular medication due to some other disease. Furthermore, the chance of co-morbidities is rather high in psychiatric disorders (Thaipisuttikul et al., 2014; Zimmerman et al., 2002), therefore co-morbidities be need to be considered and included at some point, another argument for the multi-dimensionality in the target variable for research. However, to capture the variability introduced when taking all these possibilities into account, much larger data sets are needed. The TDBRAIN and B-SNIP collections are data sets that aim in this direction by including several psychiatric diseases by design. At least, instead of neglecting additional diagnostic information, those should be thoroughly recorded like, e.g. in the CAV data set.

Providing a detailed description about the research sample is necessary to enable replicability of the research, a cornerstone of good scientific practice. While there are limits to the level of detail that can reasonably be provided about the participants, some papers do not provide the bare minimum. We found one paper where we could not determine the overall sample size and four more without information about the gender ratio. Statistics about age was missing in five studies. One study was missing the inclusion criteria for the MDD group and eight studies for the inclusion in the HC group. Finally, ten studies did not provide any information about drug-status of any of their participants.

Given the influence these factors have on EEG-biomarker for MDD, these studies do not add useful findings to the research topic.

## 5 Conclusion

Detailed demographic, diagnostic, psychological, social, and behavioral characteristics of the study population should be collected and reported, as well as considered in the analyses. Moreover, the variability in study population should be taken into account when conducting reviews or meta-analyses.

Larger data sets that are more diverse by design are needed, ideally publicly available. The size of the data set comprises on the one hand the number of participants but also the information gathered about the participants. This review shows that there are already several well-curated data sets publicly available, and used in studies. However, the possibilities given with these sets to replicate results based on one’s own data and therefore strengthen the generalizability of the research findings are still widely neglected.

Rather than a dichotomous separation in MDD and HC groups, the target variable should be either a continuous metric of depression severity or even better a multi-dimensional characterization of different (patho-)psychological dimensions. These target variables should ideally cover the whole spectrum without gaps or missing extrema.

Considering all these issues should bring clinical researchers a step closer to enable robust decision support for MDD diagnostics. The support procedure might then provide a fine-grained, multi-dimensional characterization of the patient beyond the label MDD. It enables the discovery of co-morbidities and provide a better demarcation to other psychiatric disorders. Furthermore, this patient stratification might help in increasing treatment success. Finally, this approach is well-suited to shed light the physiological underpinnings of the disorder.

## 7 Author Contributions

RM: Data curation, Writing – original draft, Writing – review & editing; AR: Conceptualization, Funding acquisition, Supervision, Writing – original draft, Writing – review & editing

## 10 Data Availability Statement

The data sets CAV (Cavanagh et al., 2019) and MODMA (Cai et al., 2022) that were used for the additional analyses in this study can be found on the PRED+CT website (http://predict.cs.unm.edu/) and in the MODMA repository (https://modma.lzu.edu.cn/data/index/).

